# Implementing the mhGAP-HIG: The process and outcome of supervising trained primary health care workers in Khyber Pakhtunkhwa, Pakistan

**DOI:** 10.1101/2024.11.29.24317879

**Authors:** Asma Humayun, Arooj Najmussaqib, Noor ul Ain Muneeb

## Abstract

**Background:** The province of Khyber Pakhtunkhwa (KP), grappling with frequent humanitarian and conflict challenges, faces significant gaps in mental health services marked by limited resources and inequitable distribution of services. To strengthen these services in nine districts in the province, 105 PHCWs were trained to identify and treat psychological conditions and were subsequently supervised for three months. This study examined the efficacy of remote supervision and analyzed the clinical data gathered during the supervision period.

**Methods:** A mixed-method approach was used to collect clinical data during supervision. Supervision covered assessment, management (including pharmacological and psychosocial interventions), and referral needs in all cases. Both qualitative and quantitative feedback were analyzed. Additionally, clinical data were examined to identify reported stressors and clinical presentations.

**Results:** Out of 105 registered trainees, 53 (50.34%) participants (including 38 PCPs and 15 CPs) submitted 413 cases through the application during three months of supervision following the initial training. The most frequently reported condition was depression (56.9%). Commonly reported stressors include health challenges or caregiver burden, marital or domestic challenges, bereavement, and socio-economic difficulties. Supervision was crucial in adjusting diagnoses in nearly a quarter of cases and management plans in 38.25% of cases. Participants expressed a preference for remote supervision and found it beneficial for assessment/diagnosis (61.1%), management interventions (72.2%), and referral guidance (44.4%).

**Conclusion:** Effective capacity building of PHCWs depends on remote supervision for an extended period, continuous monitoring of assessment and intervention skills, and the establishment of structured referral pathways. The collection of clinical data is crucial for improving the training programs. Systematic support from provincial governments is essential to scale up this initiative.

---

The province of Khyber Pakhtunkhwa (KP) is in the northwestern region of Pakistan and shares an international border with Afghanistan to the west. As a result, the province has been facing geopolitical conflicts and violence for decades. The province’s susceptibility has intensified because it hosts more than 58% of the total Afghan refugees in Pakistan (1) and has also suffered an economic decline due to the impact of climate change, such as floods in 2022, where 17 out of 37 districts were affected (2,3).

Unfortunately, Pakistan ranks 154^th^ among 195 countries in terms of healthcare access and quality index (4). Due to the constraints of the health budget, the allocation designated for mental health is only a small fraction of the total (5). The implementation of mental health plans is consistently impeded by a lack of human and financial resources (6). Confronted by recurring humanitarian challenges, the lowest gains in life expectancy have been reported in the provinces of KP and Baluchistan between 1990 and 2019 compared to the rest of the country (7). The gaps in mental health services in KP are also marked by limited resources and inequitable distribution of services, which predominantly centers on a biomedical approach (8).

In 2023, in collaboration with the health department in KP and the International Medical Corps (IMC), a pilot project was conducted to strengthen mental healthcare services for Afghan refugees and host communities in nine districts (Chitral, Haripur, Kohat, Lower Dir, Mansehra, Mardan, Nowshera, Peshawar, and Swabi). Using the developed tools, 105 primary health care workers (PHCWs), including primary care physicians (PCP) and clinical psychologists (CP), were trained in six 5-day workshops (9). After the training, significant improvements in the participants’ knowledge to identify and treat mental health conditions were noted. The feedback and reflections of the participants also highlight the strengths and gaps of the training process. As part of this initiative, trained PHCWs were supervised for at least three months.

Following the training process (9), this paper discusses the process of remote supervision and the analysis of clinical data collected through the process of supervision.

## Use of digital technology to provide supervision

Past initiatives to build the capacity of PHCWs in Pakistan emphasized initial training, but the planning for a supervisory mechanism was inadequate (10,11). Some of the challenges in providing post-training supervision in the context of KP include a severe dearth and inequitable distribution of existing mental health services, identifying suitable trainers who are motivated to strengthen mental health services in primary care, and the lack of incentives for supervisors and logistical challenges of providing supervision in geographically remote areas (8).

Digital technology is known to help mobilize mental health services in remote and conflict-affected areas where either healthcare facilities do not exist or are difficult to access (12–14). Peer support and remote supervision are recommended when face-to-face supervision is not feasible in humanitarian settings (15). Mobile phones have also provided access to mental health services, which is otherwise not possible offered after the Iraq War and among Syrian refugees (13,14). It is also known that technological solutions such as audio recording and WhatsApp groups can help promote a better quality of supervision when there is an inadequate number of specialists to provide supervision (16). In resource-poor settings, such as KP, technology-assisted training in primary healthcare systems has already shown successful outcomes in delivering psychosocial interventions (17). The trend of relying on digital tools to strengthen routine services in LMICs has also been increasing (18,19).

## The mhGAP-HIG-PK mobile application

Following the recommendations of the World Health Organization (2021), an evidence-based, digital, and scalable MHPSS model was developed by the Ministry of Planning, Development & Special Initiative (MoPD&SI) in 2021 to strengthen the existing mental healthcare system (21). As part of this work, adapted from the mhGAP Humanitarian Intervention Guidelines (22), capacity-building tools (mhGAP-HIG-PK) were also developed for Pakistan (23). These include a specifically designed mobile application that provides a complete reference to the guidance and provision for case-based supervision. All conditions were indexed and color-coded as in mhGAP-HIG on the homepage. After a module is selected, the assessment and management protocols are presented in a single-page series with a proceed button, indicating when the end of the algorithm has been reached. There are choices to select the language (English or Urdu) for the clinical tools (interview questions to elicit history and symptoms, step-by-step examinations, and intervention techniques) alongside the protocols.

Compared to other smartphone-based applications being used for remote supervision (19,24,25), our application does not have a decision-making function and requires less documentation, less storage, and low bandwidth. Most app functionality works even without the internet. The cases that are entered are saved in offline storage and synchronized with the server whenever the user connects to the internet. In addition to an easy-to-use design with limited text entry, it allows the user to request urgent supervision to discuss cases and make clinical decisions.

## Method

### Setting and Study Design

mhGAP-HIG-PK training was conducted in nine districts (Chitral, Haripur, Kohat, Lower Dir, Mansehra, Mardan, Nowshera, Peshawar, and Swabi) of Khyber Pakhtunkhwa. A mixed-methods approach was employed for the collection of clinical data during supervision through an online web portal. This study also explored the perceptions and experiences of PHCWs in utilizing remote supervision, including its advantages, challenges, and recommendations.

The study participants were PHCWs who had undergone training in mhGAP-HIG-PK. The sample included 105 participants (64% males) working in the nine districts mentioned above, of which 70 were PCPs and 31 were CPs. All participants were registered on the MHPSS web portal, which is a platform designed for the assessment and reporting of cases for supervision. Nearly half of the trainees (51.5%) reported clinical cases used for analysis in this study. Amongst the other half, some worked as hospital administrators and therefore did not offer clinical services; others could not participate because of their professional commitments, and a few remained unresponsive post-training without offering any explanation. Very few cases were reported from Mansehra; therefore, the data were merged with Haripur for analysis.

### Process of supervision

Supervision was provided by the master trainer alongside a team of 7 trainers (4 psychiatrists, and 3 clinical psychologists) voluntarily through case-based discussions. Due to logistic and resource constraints, remote supervision (through a mobile application) and peer support (through WhatsApp groups) were provided for three consecutive months post-training.

Trained PHCWs were requested to submit at least 10 cases to qualify for training certification. After seeking permission from the patients, they were required to record brief clinical descriptions in the application. The patients’ contact details were optional. After submission of the case, it was reviewed by the supervisors on the dashboard, who either sent a message or called the reporter to discuss the case within three days in routine and within 24 hours if it was marked as urgent. The participants were supervised for assessment and management, including both pharmacological and psychosocial interventions and the need for referral. Based on the available clinical information and mhGAP protocols, the performance of the PHCWs was evaluated for case-detection accuracy and choice of intervention. Lastly, the supervisors recorded the ‘outcome’ of the discussion on the portal, which included brief comments about additional assessment, modified diagnosis, and any advice offered for intervention, including referral.

PHCWs who had submitted cases were invited to provide feedback on the process and quality of supervision. A brief questionnaire consisting of 10 items was adapted from an interview guide administered as part of the Emilia Project (19). The set of questions aimed to evaluate different aspects, including professional activities, technology usage, Internet accessibility, and the degree to which the mhGAP guide and application supported remote supervision during the process.

In addition to case-based supervision through the application, six WhatsApp groups were established following each training workshop, which served as peer support groups. The objective was to facilitate the process of training and supervision, technical or logistics support, share resources, encourage peer support, and align with the strategy of the Communities of Practice (COP) (26). Each group comprised trainees, trainers, and representatives from IMC. These groups were instrumental in offering post-training educational, supervisory, and mentoring support, thereby ensuring the practical application of acquired knowledge and skills by the participants, and were intact at the time of writing this paper. They were encouraged to share cases and foster peer interactions and collaborative learning.

### Ethical considerations

This study was conducted as part of the Mental Health and Psychosocial Support Project, approved by the Ministry of Planning, Development & Special Initiatives in compliance with ethical standards and consent protocols under letter no. 6(262) HPC/2020. All the collected data were anonymized and digitally encoded. Access to personal information was restricted to the research team. Verbal consent was obtained from patients to use their clinical data for research purposes. Furthermore, all WhatsApp groups were moderated to guarantee mutual respect and to maintain the confidentiality of clinical data shared during case discussions.

### Data analysis

Data analysis was performed in various steps. The initial objective was to analyze the supervision process using a thorough quantitative analysis to assess both the reported information and input provided during supervision. We used descriptive analyses such as calculating the mean, percentage, and grouping based on demographic information.

In addition, we conducted a comprehensive hybrid theme analysis that combined qualitative and quantitative methodologies to examine the reported stressors associated with mental health conditions MHCs). For the thematic analysis, two independent researchers identified physical symptoms and stressors reported in the data of clients. After comprehensive familiarization, each researcher reviewed the data to thoroughly understand the content and context. Following familiarization, the researchers independently generated initial codes for possible stressors. These codes were then collated into potential themes reflecting the broader patterns of stressors that emerged across the data. To ensure the robustness of our thematic analysis, the identified themes were reviewed by a third researcher to ensure that the identified themes adequately represented codes and data representation. Finally, the definition and naming of the themes were finalized by comparing thematic descriptions and discussions between the researchers to achieve consensus. Once the themes were finalized, a quantitative content analysis was performed. This included counting and analyzing the identified themes within the qualitative data. A similar process was repeated to analyze the reported somatic symptoms.

## Results

In this section, we present the details of the total number of reported cases, including the demographic profile of individuals presenting with MHCs to primary care centers, their clinical presentations, sources of stress, and reported mental health conditions. We also present the outcome of the supervision process in terms of the accuracy of identification of MHCs and interventions offered in accordance with mhGAP-HIG protocols.

The purpose of this section is to evaluate clinical cases reported by PHWCs for supervision, identify the strengths and gaps of training to help PHCWs identify and manage mental health conditions, analyze the feedback from the participants, and assess the efficacy of case-based remote supervision.

### a. Reported cases during supervision

Out of 105 registered trainees, 53 (50.34%) participants (including 38 PCPs and 15 CPs) submitted 413 cases through the application during the supervision period of three months post-training. Urgent supervision was sought for only 12 patients. A total of 26 participants reported 10 or more cases; the diagnostic range varied between one and eight MHCs. The highest number of cases was reported in Mardan (23%), followed by Lower Dir (18%), Chitral (17%), and Peshawar (14%). However, reporters from Haripur, Nowshera, and Swabi had relatively lower figures, ranging from 9% to 10%.

Of the reported cases, 68.28% (282) were women and 31.72% (131) were men. More than 70% of the reported cases (n=300) were between the ages of 21 and 50 years, with demographic cohorts of 21-30 (31%), 31-40 (25%), and 41-50 (16.2%). There were 69 cases (16.7%) under the age of 20 years, while only 11 cases (2.7%) were reported over the age of 60 years.

### b. Reported clinical presentations

This section includes an analysis of frequently reported clinical presentations and comorbidities. The most commonly reported presentations were symptoms of depression and somatic symptoms.

For depression, the symptoms reported (with frequency) were as follows: Loss of interest or ability to enjoy (67), sleep problems (61), loss of appetite (37), low mood (36), reduced energy or fatigue (36), negative thoughts or hopelessness (29), and a lack of concentration (28). Other reported symptoms include changes in body weight, reduced libido or sexual problems, and current thoughts or acts of self-harm. Dissociative symptoms, urinary incontinence, and nocturnal enuresis have also been reported.

Analysis of the data showed that somatic symptoms were frequently reported. Table 1 shows the thematic analysis and frequency of reported somatic symptoms. Neurological symptoms were the most common, followed by gastrointestinal complaints and musculoskeletal and cardiac symptoms.

**Table 1:** Thematic analysis of reported somatic symptoms with frequency.

| Category | Frequency |
| --- | --- |
| **Neurological** | 55 |
| Headache (29); Dizziness (12); Numbness in limbs (7); Fainting (5); Jaw lock (1); Jerky movements (1). |  |
| **Gastro-intestinal** | 53 |
| Abdominal/epigastric pain or discomfort (38); Dyspepsia (10); Nausea (3); Constipation (2). |  |
| **Musculoskeletal** | 39 |
| Generalized body aches & pain (26); Specific pain e.g., back, shoulder, neck (13). |  |
| **Cardiovascular** | 32 |
| Palpitations (23); Chest pain (7); Heavy heart (2). |  |
| **Respiratory** (Shortness of breath). | 8 |
| **Miscellaneous** | 8 |
| Excessive salivation (1); Nasal bleeding and foaming at mouth (1); Ringing in ears (1); Nightfall (1); Unspecified (4). |  |
| **Total** | **195** |

Comorbid conditions were reported by 57 patients (13.8%). Unsurprisingly, hypertension and diabetes have been frequently reported. Other comorbidities included urinary tract infections, anemia, Parkinson’s disease, asthma, and contact dermatitis.

### c. Reported stressors

PHCWs have reported a range of current and past sources of stress in clinical cases. The thematic analysis of stressors provided valuable insights into the social and environmental factors of individuals presenting with MHCs (Table 2). A total of 442 stressors were categorized based on the available data.

**Table 2:** Thematic analysis of reported stressors (with frequencies)

|  | Stressors<br>(sub-themes) | No of<br>cases |
| --- | --- | --- |
| 1. | **Socio-economic** | 72 |
| 2 | **Crisis/Trauma**<br>Traumatic event (21); Disputes (14); Displacement (7) | 42 |
| 3 | **Bereavement**<br>Death of loved ones (parent, partner, sibling, child) | 82 |
| 4 | **Marital**<br>Marital discord/ divorce/forced marriage (24); Domestic violence (24); Lack of support (46) | 94 |
| 5 | **Health**<br>Own health (36); Fertility related (10); Family's health (15); Carer's burden (38) | 99 |
| 6 | **Other**<br>Family (38); Occupational (15) | 53 |
|  | **Total** | **442** |

Stress factors related to one’s health, including caregiver burden and family health, were the most documented (99 cases). Carers’ burdens included childcare or looking after an unwell family member. Issues related to fertility were specifically mentioned for married women. These stressors have a dual influence on patients’ mental health, both directly through physiological and psychological strains and indirectly through the socio-economic burdens imposed by healthcare costs and caring responsibilities. Factors related to marital stress were also frequently reported in women (94 cases). This category includes cases of lack of support, marital discord, divorce, forced marriage, and domestic violence. Bereavement as a stressor was noted in 82 cases, predominantly due to the death of a close family member. Many people have reported socio-economic stressors, trauma-related crises, ongoing disputes, or displacement. Some cases also reported challenges related to their families, education, or occupation.

### d. Outcome of supervision: Assessment

Of the 413 cases, the reported diagnosis was adjusted for 102 cases (24.70%) after supervision. Among these, there were 24 (5.81%) cases in which the diagnosis was inconclusive, that is, either due to an incomplete assessment or did not merit a diagnosis based on the available information. Table 1 presents the frequency of reported MHCs enlisted according to the mhGAP-HIG, both reported and adjusted after supervision. The MHCs in which the frequency increased after supervision included Dep (15 cases), acute stress (5 cases), grief (7 cases), and conditions that were over-reported and did not meet the criteria following the mhGAP-HIG included PTSD (15), psychosis (8), epilepsy (2), and SUB (6).

Following supervision, DEP was the most commonly reported condition, with 235 cases (56.9%) in 174 women and 61 men, ranging from 11 to 50 years of age. This was followed by ACU and GRI (9% each). The least reported conditions were PSY (1%), SUB, PTSD, and suicide (2% each).

Table 3 shows the reported and adjusted diagnoses and frequency of reported MHCs for each gender. DEP (34), GRI (34), and OTH (19) were much more commonly reported in women, whereas SUB was predominantly reported in men (6), with five cases recorded in young men (21-30 years). ACU was nearly equally reported in both men and women (17 and 19 cases, respectively).

**Table 3:** Reported and adjusted (post-supervision) frequency of MHCs.

| Conditions | Reported |  | Adjusted |  | Males |  | Females |  |
| --- | --- | --- | --- | --- | --- | --- | --- | --- |
|  | F | % | F | (%) | F | % | F | (%) |
| ACU | 31 | 7.51% | 36 | 8.72% | 17 | 4.12% | 19 | 4.60% |
| GRI | 31 | 7.51% | 38 | 9.20% | 4 | 0.97% | 34 | 8.23% |
| DEP | 220 | 53.27% | 235 | 56.90% | 61 | 14.77% | 174 | 42.13% |
| PTSD | 22 | 5.33% | 7 | 1.70% | 4 | 0.97% | 3 | 0.73% |
| PSY | 13 | 3.15% | 5 | 1.21% | 2 | 0.48% | 3 | 0.73% |
| EPI | 20 | 4.84% | 18 | 4.36% | 10 | 2.42% | 8 | 1.94% |
| ID | 10 | 2.42% | 10 | 2.42% | 6 | 1.45% | 4 | 0.97% |
| SUB | 13 | 3.15% | 7 | 1.69% | 6 | 1.45% | 1 | 0.24% |
| SUI | 10 | 2.42% | 8 | 1.94% | 3 | 0.73% | 5 | 1.21% |
| OTH | 43 | 10.41% | 25 | 6.05% | 6 | 1.45% | 19 | 4.60% |
| Incomplete Assessment | 0 | 0% | 24 | 5.82% | 12 | 2.91% | 12 | 2.91% |
|  | **413** |  | **413** |  | **131** | **31.72%** | **282** | **68.28%** |

*Note.* The full forms were as per mhGAP-HIG. ACU (acute Stress); GRI (grief); DEP (moderate-severe depressive disorder); PTSD (post-traumatic stress disorder); PSY (psychosis); EPI (epilepsy/seizures); ID (intellectual disability); SUB (harmful use of alcohol & drugs); SUI (suicide); OTH (other significant mental health complaints)

40% of ID cases, 25% of SUI cases, 29% of SUB cases, 20% of DEP cases, and 20% of PSY cases were reported in Chitral, a district without any psychiatric services and noted for high reports of suicide rates. Similarly, PHCWs from Lower Dir, another district without any psychiatric services, reported 40% PSY cases, 34% GRI cases, and 31% ACU cases. Furthermore, 43% of SUB and 43% of PTSD cases were reported in Peshawar, the capital of the province. Three out of eight suicide cases were reported in young people between 11-20 years.

### e. Outcome of supervision: Interventions

In addition to guiding assessment, supervision was offered for management, encompassing both pharmacological and psychosocial interventions (PSI) and referrals. Management interventions were reported in 70.7% of cases. Supervision played an important role in modifying the management plans in 158 cases (38.25%). Following supervision, the PCPs were advised to provide additional interventions in 55 cases: pharmacological interventions in 32 cases and PSI in 23 cases. There were 12 patients who were already taking benzodiazepines, and PCPs were advised to help users reduce or discontinue the medicine.

Of the 276 psychosocial interventions reported (one case could have had more than one intervention reported), 64 interventions were reported outside the mhGAP-HIG treatment protocols. Examples of such interventions include cognitive behavioral therapy (CBT), mindfulness, anger management, coping strategies, and emotional regulation techniques. All interventions were reported by the CPs.

Prior to supervision, PHCWs only referred to 33 (8%) patients. Following supervision, this figure increased to 54 (13%) as seven PCPs and 14 CPs were advised to refer the patient to a psychiatrist (35) for additional evaluation, a mhGAP-trained PCP (13) for the prescription of an antidepressant, a neurologist (2), and one each to a CP, physiotherapist, speech therapist, and ophthalmologist, respectively.

### f. Feedback from the participants

Following supervision, feedback was received online from 36 participants. All participants had access to the internet through either a mobile device or a laptop. 89% of the PHCWs had Internet access both at their workplace and at home, 8.3% had access exclusively at home, and 2.8% had access only at work. The majority of PHCWs (72.2%) indicated that they sought supervision for psychological or pharmacological interventions, 61.1% for assessment/diagnosis, and 44.4% for referral. 58.3 Of the participants, 58.3% expressed a preference for remote supervision, whereas 41.7% reported a preference for in-person supervision. All participants evaluated supervision as satisfactory, with 55.6% rating it as very satisfactory and 44.4% reporting it as satisfactory.

During consultations, 61.1% reported using both the guide and app, 19.4% reported using only the app, and 16.7% reported using only the guide. Only 2.8% of the participants had not used an app or guide. All participants unanimously endorsed mhGAP training for both PCP and CPs.

## Discussion

For the first time in Pakistan, despite limited resources and multiple challenges, we provided supervision to trained PHCWs after mhGAP-HIG training. Without any extrinsic incentive, half of the trained PHCWs actively engaged in seeking supervision for three months. In the absence of a reliable system to document or record information at primary healthcare facilities, our digital MHPSS model helped collect relevant data that were analyzed in this study. These data provide invaluable insights into mental health care needs at the primary care level and will help strengthen our training process. The mhGAP-HIG-PK mobile application, a tool under this model, facilitated the process of remote supervision by providing a secure and instant channel of communication and record-keeping.

In this section, we discuss the effectiveness and broader implications of the supervision process following mhGAP-HIG training for PHCWs.

From their feedback, it is evident that PHCWs referred to mhGAP-HIG-PK protocols during clinical consultations. Although depression was the most frequently reported condition, it was encouraging that PHCWs reported a range of conditions across all modules during supervision. However, our analysis showed clear gaps in the ‘assessment’ offered by trained PHCWs. For example, there is a clear discrepancy between the reported psychiatric and somatic symptoms. On the one hand, the frequency of reported symptoms of depression lags behind the number of cases reported within this category, and on the other hand, somatic symptoms were much more frequently reported. It is already known that most people with mental health conditions present with physical symptoms, such as headaches, pain, palpitations, and epigastric issues (27), but it is equally likely that PHCWs tend to notice or check for bodily symptoms more than psychological symptoms.

This also means that even for a condition similar to depression, PHCWs reported it without a complete assessment. Similarly, most cases where psychosis and PTSD were initially reported did not fulfill the symptom criteria during supervision. Any oddity of behavior was misinterpreted as psychosis, and the presence of a traumatic event automatically prompted the consideration of PTSD. We also found that when there was a possibility of two conditions, depression and harmful use of substances, the PHCWs were inclined to report it as SUB and disregarded depression. Incomplete assessments were found in at least 24 cases in which PHCWs were advised to review the patients, primarily considering the risk of self-harm. This is particularly important because we know that information about self-harm or suicide attempts is usually not spontaneously offered for fear of being judged (28). More emphasis and practice were needed on ‘assessments’ of MHCs during training and supervision.

Since medical training in Pakistan promotes a biomedical approach, it was encouraging that most physicians attempted to offer PSI. The literature suggests that mhGAP training helps reduce stigma and enhance confidence in psychological interventions among primary care physicians (29). Through supervision, management plans were modified well in over a third of all cases. It was noted that PHCWs, particularly CPs, were reluctant to consider that people with depression might need medication. We also noted that CPs tended to offer anger management for people with depression.

The referral mechanism for MHCs from primary care to specialists is nonexistent in Pakistan (30). According to the annual report of the District Health Information System in Khyber Pakhtunkhwa (31), psychiatric cases are among the least frequently presented in outpatient departments (OPDs), even less frequently than those for homeopathy. This inadequate system often forces patients to either directly seek specialists for common mental disorders or turn to non-conventional practitioners, such as faith healers, Hakeem, or homeopaths for assistance (6). We also found that only a small fraction of the patients was referred to specialists by PHCWs. Furthermore, physicians typically lack the necessary training to appropriately address these symptoms (32), stigma attached to a psychiatric referral, and difficulty accessing specialist services (33). After supervision, 13% of the patients were referred. In some instances, CPs were advised to refer the person to a primary care physician trained in mhGAP, mostly when the diagnosis of depression was clear and the person needed an antidepressant. We also noticed that very few primary care physicians considered referring a person to a CP.

We analyzed 413 cases, including demographic profiles, clinical presentations, and reported stressors affecting individuals with MHCs. Two-thirds of cases reported mental health conditions (MHCs) in women where conditions such as Depression, Grief, and Others being much more common. The contribution of depressive disorders to DALYs has already been found to be substantially higher in women (34). Thematic analysis of stressors revealed that health challenges or carers’ burden, marital or domestic challenges, including the absence of supportive relationships, bereavement, and socio-economic difficulties were the most common. Domestic conflicts and socio-economic challenges have also been identified as major stressors that led to suicide in Pakistan (35). In our target districts, which host large refugee populations, many stressors arise from conflicts in their home countries or from the crisis of displacement. For instance, a middle-aged refugee presented with post-traumatic stress disorder for five years after his sibling was killed in Afghanistan. Similarly, one grandmother had been suffering from depressive symptoms following the death of her grandchildren during the Afghan War.

Unsurprisingly, over half of the cases were reported to suffer from depressive disorder, which is known to be the leading cause of mental health burden globally and in Pakistan (34, 36). However, the condition might have been overrepresented in our cases because mhGAP-HIG considers reactions to acute stress and grief within a month of the stressful event or loss, so people presenting with stress-related conditions after a month might also have been recorded as depressed by the PHCWs. In addition, people presenting with anxiety symptoms have also been reported to have depression.

Additionally, comorbidity was reported in only 14% of cases. Encouragingly, in a young lady who presented with skin rashes and was treated for contact dermatitis, PHCW explored stress and found that she had sleep disturbances, loss of appetite, and irritability. She had two daughters, faced the pressure of not having a male child, and was now worried that her husband would remarry. Globally, the prevalence of the co-occurrence of mental and physical illnesses is increasing, even in the younger population, but it is often overlooked during medical consultations (37). We feel that more emphasis needs to be placed on checking for comorbidities during training.

Our experience of providing remote supervision was the most encouraging, as the majority of our target districts lacked psychiatric services. We also received positive feedback regarding remote supervision from the trained PHCWs. Given the dearth of specialist resources in the country, remote supervision is also an opportunity to engage specialists from elsewhere in the country, including the diaspora, to help build the capacity of PHCWs. For a population of 40 million, there is no single child psychiatrist in the province of KP. In our case, during the period of supervision, a PHCW from Chitral reported the case of a child who was presented with developmental difficulties. A child psychiatrist (of Pakistani origin) working in the UK offered to supervise the PHCW to work with the family.

Online COPs have been recognized as both feasible and effective methods of supervision for building the capacity of PHCWs, especially in LMICs (38). The PHCWs were actively engaged in the WhatsApp groups, created to provide supervision and peer support, even after completing the period of supervision. Most of them worked in isolated health facilities without much opportunity for mentoring or professional development. Feedback on their cases, both from the trainers and peers, was well received and positively reinforced their motivation.

### Limitations and future directions

We learned from our experience that supervision should be provided for at least six months, as pilot trials conducted over a six-month period have been shown to be significantly effective (39). In our case, only half of the trained PHCWs were actively engaged in the supervision process. For the ongoing supervision of healthcare providers and their lasting impact, systematic support by provincial governments and policies is inevitable (40). In this regard, the selection of PHCWs and the provision of service incentives are crucial. There were a few PHCWs who were already trained as family physicians, who appeared motivated and experienced training as mhGAP trainers. These can be valuable resources for districts without psychiatric services. At present, barriers to integrating mental health into primary care include a lack of support within systems and resource allocation (41). Successful training requires districts to take full ownership with continuous monitoring and robust responses (42).

Furthermore, it is vital to develop a cohort of trainers/supervisors for the province and recognize this role as part of their career development. Remote supervision must be strengthened through in-person refresher training. We also identified some gaps in the training process and collection of clinical data. These lessons will guide us in developing our training and modifying the mhGAP-HIG-PK mobile app accordingly. Finally, referral pathways need to be more effective and structured. We were unable to follow up on the cases referred to the psychiatric service, and their feedback could also help strengthen referral pathways.

**Figure 1:**
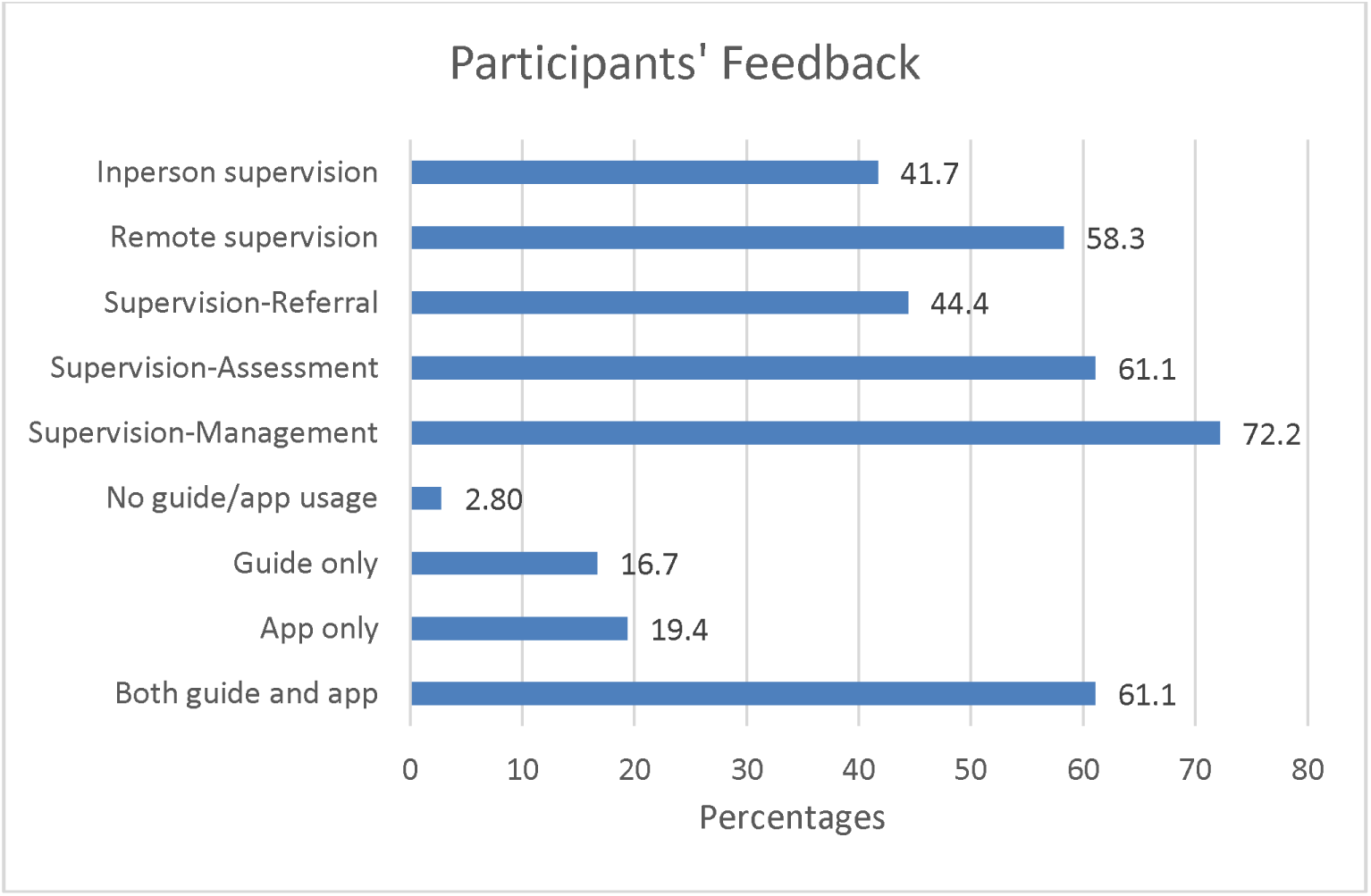
Participants’ feedback on the process and quality of supervision.

## List of abbreviations

KP: Khyber Pakhtunkhwa
UNHCR: United Nations High Commissioner for Refugees; UNHCR
UNOCHA: United Nations Office for the Coordination of Humanitarian Affairs
IMC: International Medical Corps
PHCWs: Primary Health Care Workers
PCPs: Primary Care Physicians
CPs: Clinical Psychologists
PSI: Psychological Intervention
EMRO: WHO Regional Office for the Eastern Mediterranean
LMICs: Low and Middle Income Countries
MHPSS: Mental Health and Psychosocial Support
MoPD&SI: Ministry of Planning, Development & Special Initiative
mhGAP-HIG: mental health Gap Action Program-Humanitarian Intervention Guide
mhGAP-HIG-PK: mental health Gap Action Program-Humanitarian Intervention Guide adapted for Pakistan
COP: Communities of Practice
MHCs: Mental Health Conditions
ACU: Acute stress
GRI: Grief
ID: Intellectual Disability
EPI: Epilepsy/seizures
SUB: Harmful use of substances
PSY: Psychosis
PTSD: Post-traumatic Stress Disorder
DEP: Depression
OTH: Other significant mental health complaints
CBT: Cognitive Behavioral Therapy
OPDs: Outpatient Departments

## Acknowledgements

The pilot project was undertaken in collaboration with the Department of Health, Khyber Pakhtunkhwa, and was supported by the International Medical Corps in Pakistan. We would also like to thank the primary health physicians who actively participated in supervision. Special thanks to Dr. Laiq Said Bacha and Dr. Muhammad Irshad (Lower Dir); Dr. Jamshed Khan (Mardan); Dr Sami ud Din and Dr Shafiuddin (Chitral); and Dr Said Murtaza (Haripur) for their valuable contributions.

## Ethics approval and consent to participate

This study was conducted as part of the Mental Health and Psychosocial Support Project, approved by the Ministry of Planning, Development & Special Initiatives in compliance with ethical standards and consent protocols under letter no. 6(262) HPC/2020.

## Consent for publication

All authors consented to publication.

## Availability of data and materials

The data that support the findings of this study are available from the Health Section at the Ministry of Planning, Development & Special Initiatives, Government of Pakistan. Confidentiality restrictions apply to the availability of the data used under the license for the current study and so are not publicly available. However, the data can be available from the authors upon reasonable request after formal approval from the Ministry of Planning, Development & Special Initiatives, Pakistan.

## Author’s contributions

This study was conceptualized by AH, who also led the mhGAP-HIG project in KP, from which the data for this study were derived. Both AN and NM handled data curation and thematic analysis. AH and AN jointly devised the methodology. All the authors contributed to the initial drafting of the manuscript. AH reviewed and edited the final manuscript.

